# *(How) Do we theorize?* A focused mapping review and synthesis of theoretical nursing research in the German-speaking area of Europe

**DOI:** 10.1101/2022.12.21.22283767

**Authors:** Jasmin Eppel-Meichlinger, Ana Cartaxo, Theresa Clement, Julian Hirt, Martin Wallner, Hanna Mayer

## Abstract

**Introduction:** Although nursing has made significant progress in establishing as an academic discipline in the German-speaking area, discussions in the scientific community are increasing about lack of theory dynamics. Theories in nursing provide foundational knowledge that guides actions in nursing practice, research, and education. However, nursing research has not yet been systematically investigated against this background. Our aim was to obtain an overview of the theoretical research landscape in nursing in the German-speaking countries of Europe.

**Method:** We conducted a focused mapping review and synthesis. The focus was on nursing journals articles with an explicit theory-related aim, published between 2016 and 2021 in German or English language and with a contribution of at least one author affiliated with an institution in the German-speaking area of Europe.

**Findings:** We identified 32 out of 1419 (2%) publications in nursing journals that met the eligibility criteria. Most theoretical work involved an inductive approach (n=21), with eleven papers intending to test or further develop an existing theory. Grounded theory (n=10) and conceptual analyses (n=6) were mainly referred to as the underlying methodological approach. We identified illness experience (n=8) and self-management (n=7) as the most frequently addressed phenomena. Theory-related results were predominantly theoretical models (n=13). The level of abstraction was mainly situation specific (n=20).

**Conclusions:** The proportion of publications with a theory-related aim was low. Theory-building efforts are heterogeneous and lack a recognizable pattern. More efforts should be made to develop empirically testable theories, to revise and synthesize existing theories on a meta-level. Therefore, researcher should focus on theorizing, i.e., on how to further develop existing theories as well as to develop new theories, rather than focusing exclusively on theories as results. For this purpose, nursing researchers should be properly educated in theorizing. Nursing professors have a crucial role to defy the demands of the current research mainstream and provide appropriate structures to learn and to continually practice theoretical thinking and theorizing.

## INTRODUCTION

Nursing science is considered an empirically oriented human science with an emphasis on practice, therefore the focus of research often lays on application. Nonetheless, it is also concerned with fundamental research to (further) develop the conceptual, theoretical, and methodological foundation of the discipline. Hence, the main tasks of nursing science are theory development, method development, empirical research, and implementation of the gained knowledge into practice (Brandenburg & Dorschner, 2021).

However, nursing science in the 21^st^ century has been described as being at a crossroads, with an imbalance between empirical and theoretical or philosophical inquiry threatening the development of the profession (Grace et al., 2016; Roy, 2018). Indeed, Grace et al. (2016) have cautioned that there is “the possibility of a tilt toward the empirical at the expense of the other two.” A similar observation is being discussed regarding the status of nursing theory construction in German-speaking countries of Europe, including Austria, Germany, and Switzerland, which has arguably come to a halt as a result of the strive for evidence-based nursing (Balzer, 2012; Meyer, 2010; Moers et al., 2011). In their critical essay, Moers et al. (2011) discussed the sparse theory development in Germany and noted that a gap had opened between theory development and empirical research in nursing in recent years. As theory building is defined as an indicator for the intellectual condition of a discipline, the authors expressed increasing concern about the situation in German nursing science. According to their observation, this also applied to neighboring countries, including Austria and Switzerland (Moers et al., 2011). Similar concerns had been raised earlier and continue to be addressed since the early 2000s. In 2002 Schaeffer reflected on current trends and challenges in nursing research and claimed that the state of theory development should be pushed to promote the scientific foundation of nursing. Two years later, Stemmer (2004) stated that the theory discussion has largely been abandoned and that a detachment of research from theory seems to be emerging in the development of nursing science. In 2009, Schaeffer (2009) reiterated her call for a revival of theoretical discourse, suggesting to consolidate the numerous incoherent theoretical perspectives. Moreover, Schmidli-Bless and Ricka (2011) observed a decline in the relevance of nursing theories, and Schrems (2011) noted that theory development in nursing science in German-speaking countries remains a stepchild. The existing critique culminated in the essay by Moers et al. (2011), which has kindled recurring critical debate ever since. Still in 2018, Bartholomeyczik (2018) lamented that a theory of nursing or even a theory of nursing interventions is lacking or has not been developed, and one year later Brandenburg (2019), as the situation had remained unchanged, diagnosed a widespread abstinence from theory in nursing science.

Moers et al. (2011) identified two main causes of this predicament: the increasing demands and pressures of academia, with researchers being “too busy to think”, i.e., too busy to construct theory, as well as a shift in focus from theory development to evidence-based nursing and intervention effectiveness research. Contributing factors include the fragmentation of higher education (Mayer, 2019) the “science circus” (Bartholomeyczik, 2018) with its tendency towards regulation, and the pressure to publish that scholars are exposed to (Bartholomeyczik, 2018; Stemmer, 2004).

The relevance of theories and their (further) development in nursing is based on the understanding that theories form a core body of knowledge related to significant issues in a field and set the thematic boundaries of a discipline (Meleis, 2018). Theories are central to scientific understanding because they help to gain a deeper understanding of phenomena by looking behind observed patterns and trying to explain them. In this way, theories allow us to identify relationships between phenomena that would otherwise remain unidentified. Furthermore, they make statements about the underlying condition or underlying causes of phenomena, which should be prevented or promoted. Theories are useful, for example, in developing and evaluating effective health interventions (Risjord, 2019). Not only theory and practice, but also theory and research are closely and interactively related. Scientific discoveries are particularly useful when they are organised as coherent wholes, such as theories. The information theories provide help in the conduct of studies by providing orientation regarding the context, efficiency and structure of the research design (Bradbury-Jones et al., 2022). Theory building represents the heart of the scientific process of any discipline in its own right (Schrems, 2011) and can thus be seen as an indicator of maturity (Mayer, 2019). The progress of a discipline can be measured by the scope and quality of its theories and the extent to which its scientific community engages with, uses and develops its theories (Gasser et al., 2012).

Nursing theories are defined as an organised, coherent, and systematic articulation of a set of propositions to represent aspects of reality for the purpose of describing, explaining, and predicting reactions, events, situations, conditions, or relationships (Meleis, 2018). They can be classified according to different criteria, such as their level of abstraction, their goal orientation, type of construct, and based on the phenomena to which they refer. Meleis (2018) distinguishes descriptive from prescriptive goal orientations. The former describes relationships between phenomena, whereas the latter refers to nursing therapeutics and the outcomes of interventions. The level of abstraction can range from high to low. Grand theories are the highest in abstraction, reflect the broadest scope and provide relationships between many abstract concepts. Middle-range theories address specific phenomena or concepts and reflect practice. Situation-specific theories focus on specific nursing phenomena and are limited to specific populations or to a particular field of practice (Meleis, 2018).

Theory building is understood as a dynamic process in which theories should be continuously developed, tested, and modified according to research findings. Conducting empirical research is therefore crucial for theory dynamics in nursing (Meleis, 2018). Thus, theory development comes in many shapes and forms and includes a range of research practices, with some being more, and some being less explicit regarding their theoretical intent or utility. Jaccard and Jacoby (2020) identified 16 different ways of making a theoretical contribution (Table 1) (Jaccard & Jacoby, 2020).

**Table 1:** 16 ways of making a theoretical contribution (Jaccard & Jacoby, 2020)

|  |  |
| --- | --- |
| 1 | Clarify, refine, or challenge the conceptualization of a variable/concept |
| 2 | Create a new variable or constellation of variables that are of theoretical interest |
| 3 | Add one or more explanatory variables for an outcome that have not been considered in prior work |
| 4 | Identify the mechanisms or intervening processes responsible for an effect of one variable on another |
| 5 | Identify the boundary conditions of an effect of one variable on another |
| 6 | Identify variables that moderate the effect of one variable on another |
| 7 | Extend an existing theory or idea to a new context |
| 8 | Identify nuanced functional forms of relationships |
| 9 | Identify unenunciated/unanticipated consequences of an event |
| 10 | Enrich and deepen understanding of established quantitative associations |
| 11 | Develop typologies/taxonomies |
| 12 | Import or apply grand theories and frameworks from other disciplines |
| 13 | Synthesize multiple theories into a unified framework |
| 14 | Develop a theory of measurement |
| 15 | Pit opposing theoretical explanations against one another |
| 16 | Propose alternative explanations to established phenomena |

Against the background of the discussion about sparse theory development in nursing of the German-speaking area of Europe, there was a need for an overview of the actual state of theoretical nursing research in this region. Thereby we intended to answer the questions of *whether and how we theorize* and subsequently to enable taking concerted action to reinvigorate theory discussion and development in nursing. To the best of our knowledge, no systematic assessment has yet been conducted for this geographic region to provide an overview of theoretical nursing research. Therefore, our aim was to map the landscape of theoretical nursing research by authors affiliated with an institution from a German-speaking country in Europe (Austria, Germany, Liechtenstein, German-speaking area of Switzerland). We aimed to describe the characteristics of theoretical nursing research and to contribute to future theory dynamics in nursing science of the German-speaking area by commenting on the current knowledge production of the nursing discipline.

## DESIGN

We conducted a focused mapping review and synthesis (FMRS) according to Bradbury-Jones et al. (2019) to address theoretical nursing research and its methodological characteristics in the German-speaking area of Europe (Austria, Germany, Liechtenstein, German-speaking area of Switzerland). There are three main features characterizing the methodology of FMRS which allowed us to (i) focus on a defined area of knowledge and knowledge production rather than the state of evidence on effectiveness, (ii) provide a descriptive map of research, and (iii) examine the topic in a broad epistemological context (Bradbury-Jones et al., 2019). We followed the guidance of PRISMA Extension for Scoping Reviews (PRISMA-ScR) (Tricco et al., 2018) for reporting.

## MATERIALS AND METHODS

### Eligibility criteria

As we were interested in original work and aimed to exclude discussion articles, we included original research articles with one of the following aims: to 1) develop, 2) analyse or 3) expand a theory, a concept, or a model, published between 2016 and 2021 (as of March 29) in a national or international peer-reviewed nursing journal. The primary criterion for the identification of theoretical nursing research was operationalized and further developed during several discursive rounds with all authors until a common understanding was reached. This need for calibration is described by Bradbury-Jones et al. (2019) as frequent points of contact and deliberation among the review team in agreeing the parameters of the review in order to calibrate for shared understandings about theoretical nursing research. Articles were eligible if at least one author from the German-speaking area was affiliated, irrespective of authorship position (Table 2).

**Table 2:**
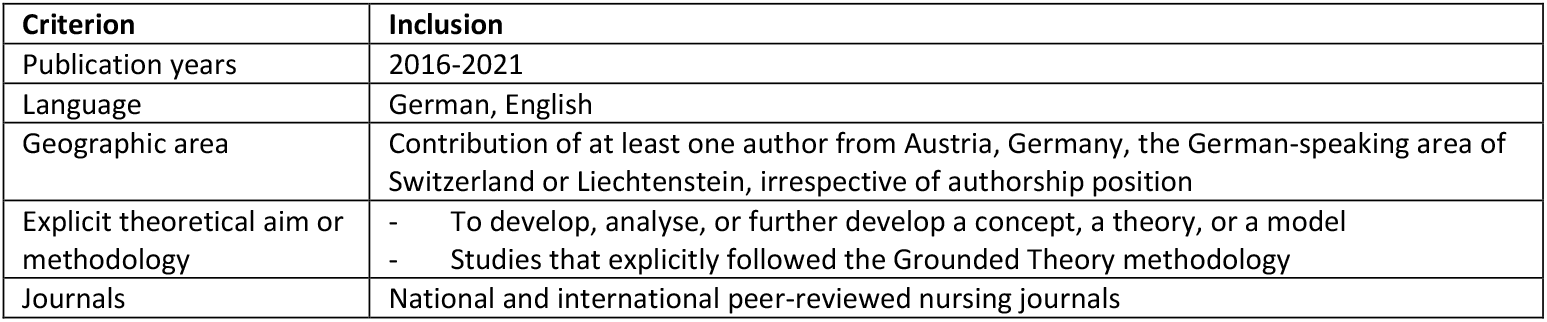
Inclusion criteria

### Information sources

#### Identification of nursing journals

We identified international peer-reviewed nursing journals assigned to the category “Nursing” in the Journal Citation Reports (JCR) (n=123, as of March 29, 2021, Appendix 1). JCR is a report on the citation impact of a defined set of journals at a given moment in time, that categorises journals using a range of bibliometric, content and subject criteria (Clarivate Analytics, 2021). Additionally, we identified three national peer-reviewed nursing journals that were not listed in JCR, by consulting the indexation overview of health science journals from the German-speaking area for the field of nursing according to Hirt et al. (2020).

#### Identification of publications

JCR journals are indexed in the Web of Science databases (Clarivate Analytics, 2021). The publications of the identified nursing journals were therefore retrieved via the database Web of Science Core Collection (WoSCC). Articles from the journals that were not listed in JCR were retrieved via LIVIVO (ZB MED, 2021).

### Study selection

By using WoSCC, publications could be automatically narrowed down to articles with at least one contribution from an author affiliated with an Institution in the German-speaking area, irrespective of the authorship position. Publications were then automatically limited to English or German language and to “Article” or “Review” as publication type. To calibrate the manual study selection process, the first 100 titles and abstracts were screened independently by four members of the author team (JEM, AC, JH, MW). This allowed the operationalization and familiarisation of the inclusion criterion “explicit theoretical aim or methodology” to be sharpened and ambiguities to be resolved prospectively. Subsequently, all remaining titles, abstracts and full texts were independently screened by two reviewers (JEM, AC). Discrepancies in the selection process between the reviewers were discussed by the research group until a consensus was found. For the study selection, we used the Systematic Reviews Web App Rayyan QCRI (Ouzzani et al., 2016).

### Data extraction and synthesis

Data extraction was performed using a standardized extraction sheet, which was developed discursively based on discussions within the author team. We considered 1) publication-related characteristics (year, journal, and country and institution of the author with an affiliation located in the German-speaking area), 2) theoretical and 3) content-related criteria, such as theory and content-related aim of the study, methodological approach, theory-related result, level of abstraction (Meleis, 2018) as well as the type of theoretical contribution (Jaccard & Jacoby, 2020). We furthermore extracted the logic of theory building (inductive or deductive), the goal orientation (Meleis, 2018), and the addressed phenomenon. Data extraction was performed by two reviewers (JEM, TC) and reviewed by each of the other. For the extraction of theoretical and content-related criteria, we used MAXQDA (VERBI Software, 2021) by descriptively coding relevant text passages to make extraction comprehensible to the second reviewer. We tabulated study characteristics, differentiating between content that was explicitly reported by the original study authors and implicit characteristics (level of abstraction, addressed phenomena, way of making a theoretical contribution) that we assessed by means of a content analysis of relevant passages of the studies, when the authors had not explicitly described their work against this background. Finally, we narratively summarized the included studies under consideration of Jaccard and Jacoby’s (18) understanding of ways to make a theoretical contribution, to give an overview of the contributions of the single theoretical nursing research publications in the German-speaking area of Europe.

## RESULTS

### Publication characteristics

#### Identification

Out of the sample with 1419 articles, we identified 32 (2%) publications with an explicit theoretical aim or methodology and the contribution of at least one author affiliated in a German-speaking country (Figure 1).

**Figure 1:**
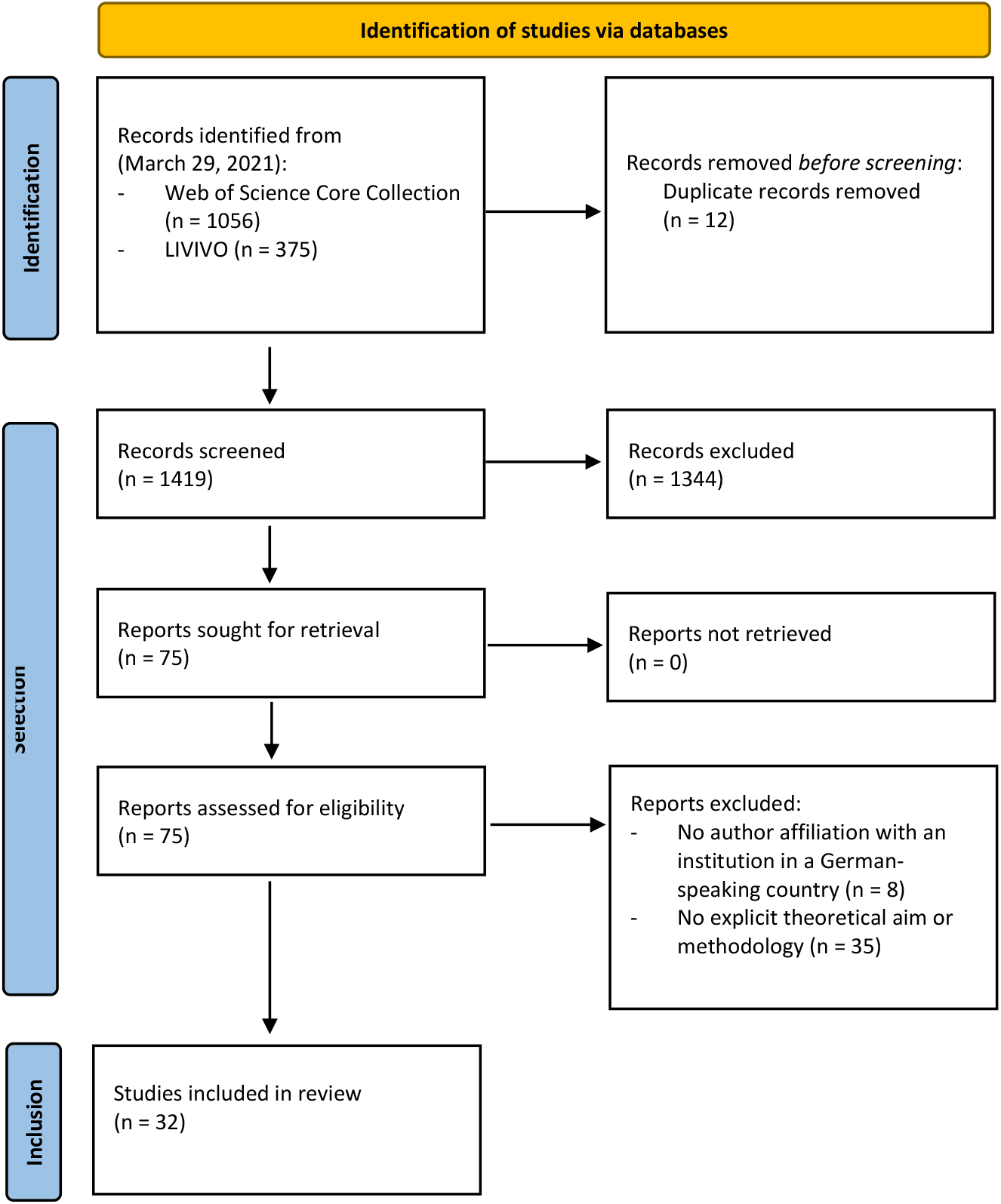
PRISMA flow diagram 2020 (Page et al., 2021) of the search and selection process

Most of theoretical nursing research was published in the years 2017 (n=8) and 2018 (n=8) (Median publication year = 2018). The annual number of publications in the focused time period ranged from three to eight. Most contributions were made by authors with a German affiliation (42%). 19 articles were published in a total of 14 different international nursing journals and 13 were published in three different journals from the German-speaking area. Contributing authors were mostly affiliated with a university (n=48, 50%). Further details are listed in Table 3.

**Table 3:** Publication characteristics of included articles; Multiple counting: *(n=41), **(n=96)

|  |  | N (%) |
| --- | --- | --- |
| Total |  | 32 (100) |
| Publication years | 2021 | 3 |
|  | 2020 | 5 |
|  | 2019 | 4 |
|  | 2018 | 8 |
|  | 2017 | 8 |
|  | 2016 | 4 |
| Affiliated geographic area* | Austria | 8 (20%) |
|  | Germany | 17 (42%) |
|  | Switzerland | 16 (38%) |
|  | Liechtenstein | 0 |
| Journals | International | 19 (59%) |
|  | Advances in Nursing Science | 2 |
|  | Applied Nursing Research | 1 |
|  | BMC Nursing | 1 |
|  | International Journal of Mental Health Nursing | 1 |
|  | International Journal of Nursing Practice | 1 |
|  | Journal of Addictions Nursing | 1 |
|  | Journal of Advanced Nursing | 2 |
|  | Journal of Clinical Nursing | 2 |
|  | Journal of Family Nursing | 1 |
|  | Journal of Nursing Scholarship | 1 |
|  | Midwifery | 1 |
|  | Nursing Philosophy | 1 |
|  | Scandinavian Journal of Caring Sciences | 3 |
|  | Women and Birth | 1 |
|  | German-speaking area | 13 (1%) |
|  | Pflege | 8 |
|  | Pflege & Gesellschaft | 3 |
|  | QuPuG: Journal für qualitative Forschung in Pflege- und Gesundheitswissenschaft | 2 |
| Type of affiliated institution** | University/University hospital | 48 (50%) |
|  | University of Applied Sciences | 34 (36%) |
|  | Clinical institution | 8 (8%) |
|  | Others | 6 (6%) |

### Theoretical characteristics

Most theoretical nursing research followed an inductive approach (n=21). Eleven articles contained the testing or further development of an existing theory, concept, or model. Grounded Theory (n=10) and conceptual analysis (n=6) were mainly referred to as the underlying methodological approach. Authors made theoretical contributions mainly by creating a new variable or constellation of variables (n=14) and by clarifying, refining, or challenging of the conceptualization of a variable or a concept (n=8). We found illness experience of patients and relatives (n=8) and self-management (n=7) as most frequently addressed phenomena. Theory-related results were predominantly theoretical models (n=13), conceptual definitions (n=6), and theories (n=6), as well as two theoretical constructs, one theoretical framework, and one nursing diagnosis. The level of abstraction of the theory-related results was largely situation-specific (n=20), ten results were middle-range. We identified three programme theories. Theoretical characteristics and their frequencies of included articles are listed in Table 4.

**Table 4:** Theoretical characteristics of included articles

|  |  | N (%) |
| --- | --- | --- |
| Total |  | 32 (100%) |
| Logic of theory building | Inductive development | 21 (66%) |
|  | Deductive development | 11 (34%) |
| Methodological approach/Methods | Grounded Theory | 10 |
|  | Concept analysis | 6 |
|  | Primary qualitative study | 5 |
|  | Meta-synthesis | 3 |
|  | Development of a programme theory | 3 |
|  | Theory synthesis | 2 |
|  | Theory analysis | 1 |
|  | Case study | 1 |
|  | Philosophical analysis | 1 |
| Addressed phenomenon | Illness experience | 8 |
|  | Self-management | 7 |
|  | Care systems | 5 |
|  | Nursing diagnostics and management | 5 |
|  | Perceptions of the discipline | 4 |
|  | Practitioners' experience | 3 |
| Theory-related result | Model | 13 |
|  | Conceptual definition | 6 |
|  | Theory | 6 |
|  | Construct | 2 |
|  | Framework | 1 |
|  | Nursing diagnosis | 1 |
|  | Others | 3 |
| Theory goal (Meleis, 2018) | Descriptive | 29 (91%) |
|  | Prescriptive | 3 (9%) |
| Level of abstraction (Meleis, 2018) | Situation-specific | 20 (58%) |
|  | Middle-range | 10 (30%) |
|  | Grand | 0 |
|  | Not assignable | 2 (12%) |
| Theoretical contribution<br>(Jaccard & Jacoby, 2020) | Creation of a new variable | 14 |
|  | Clarification, refinement, or challenge of conceptualization | 8 |
|  | Identification of mechanisms of intervening processes | 4 |
|  | Extension of an existing theory or idea to a new context | 4 |
|  | Synthesis of multiple theories into a unified framework | 2 |

Data extraction of the included studies identified as theoretical nursing research is provided in Table 5.

**Table 5:**
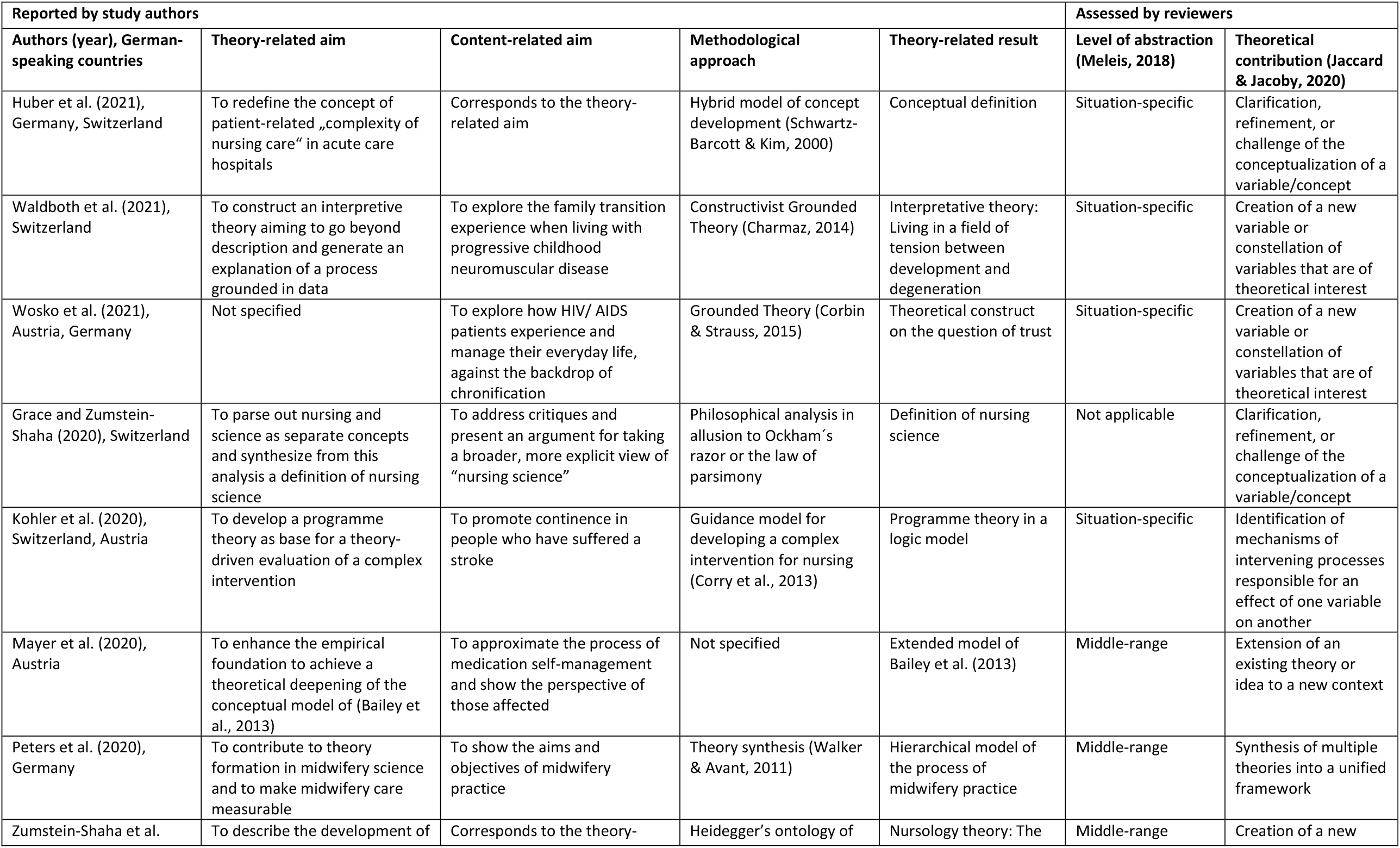
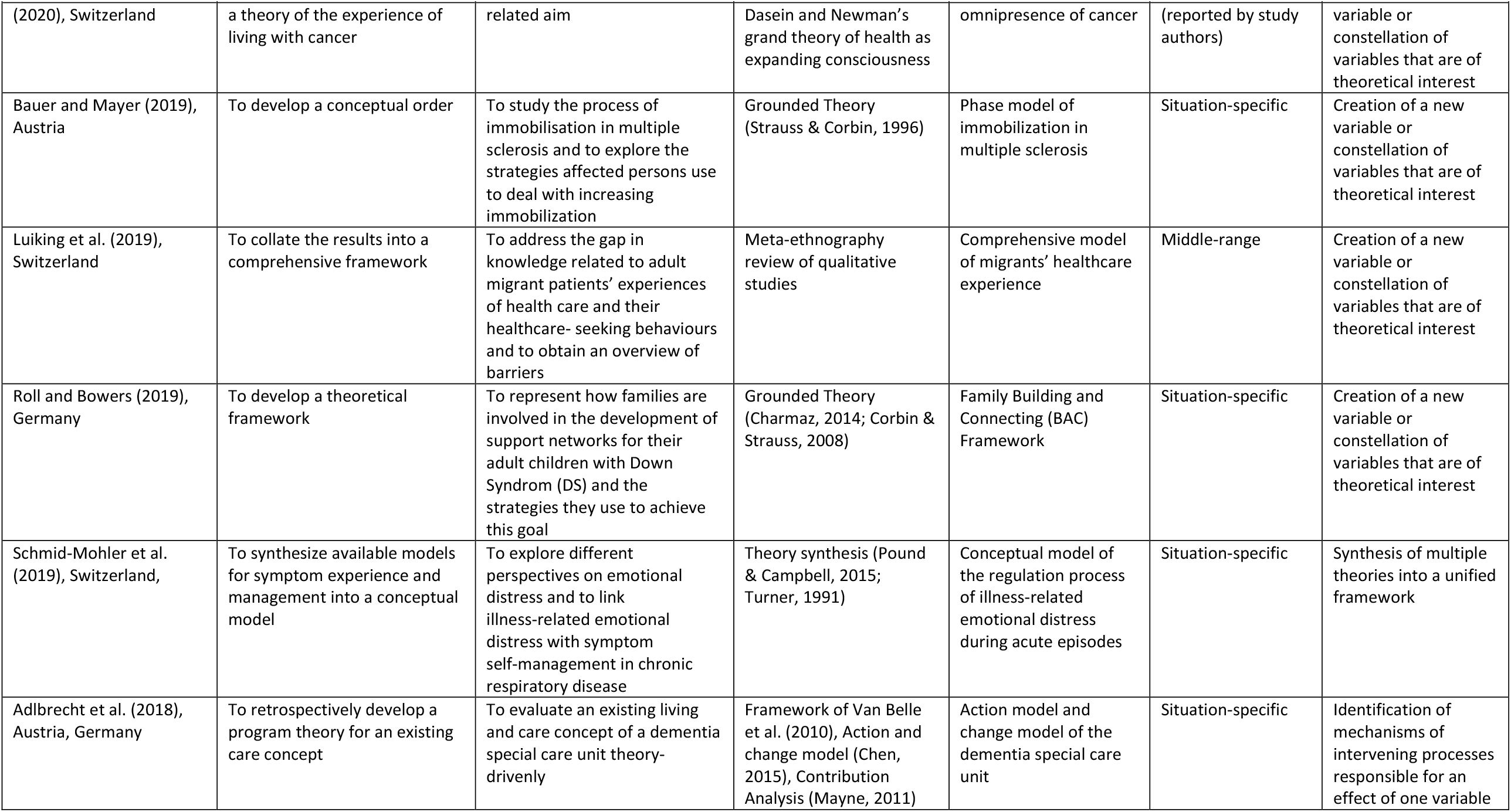
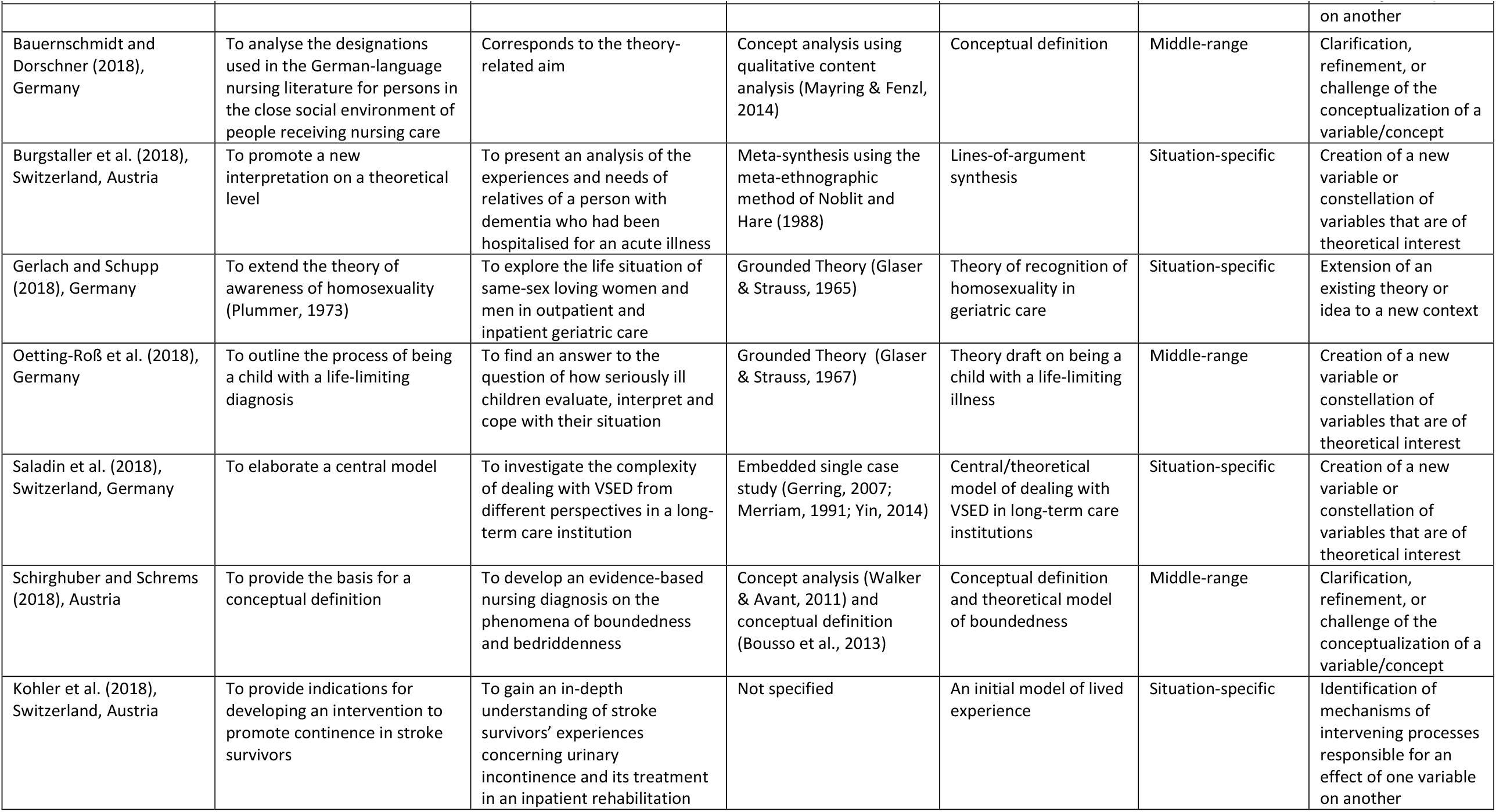
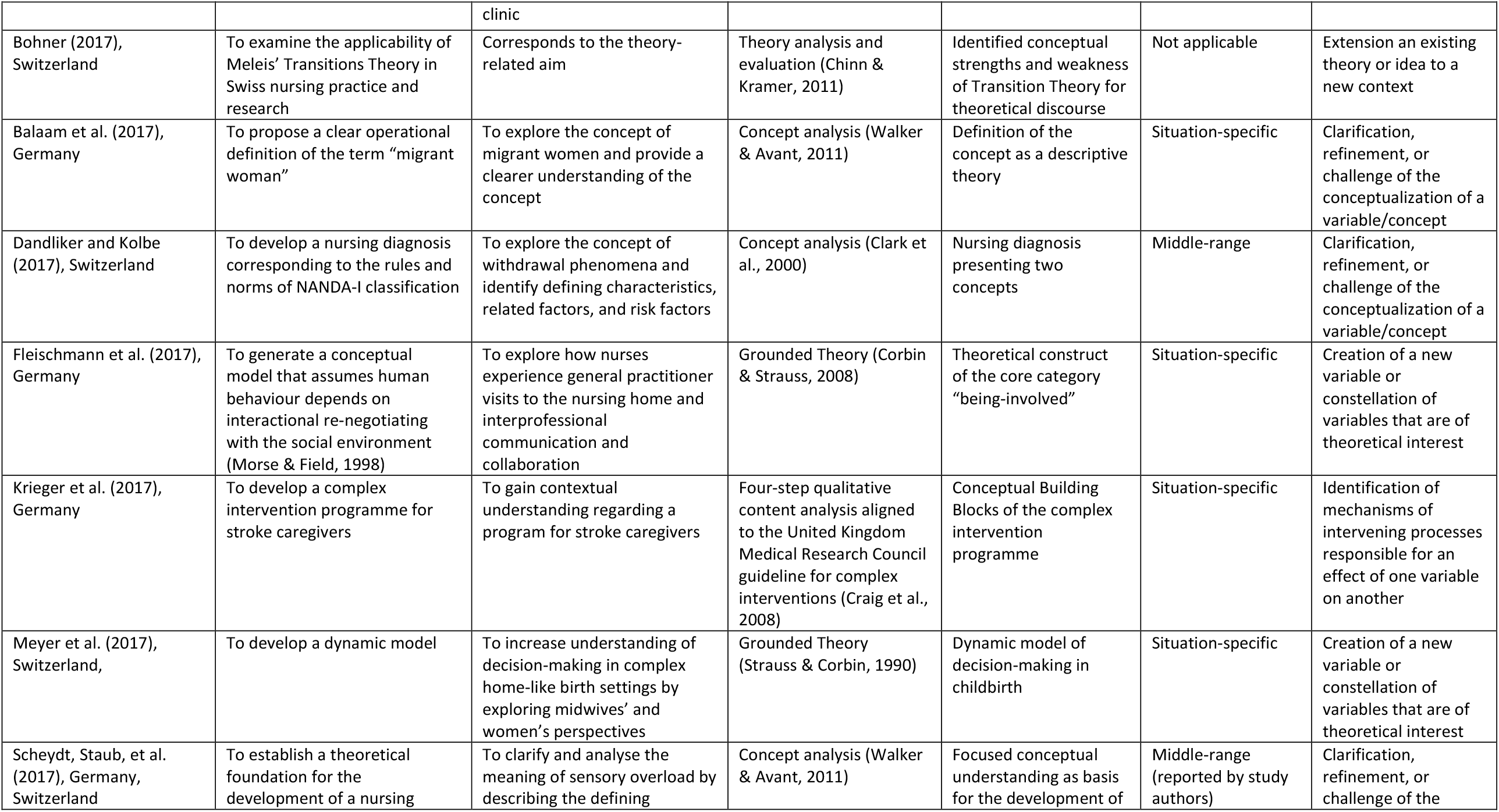
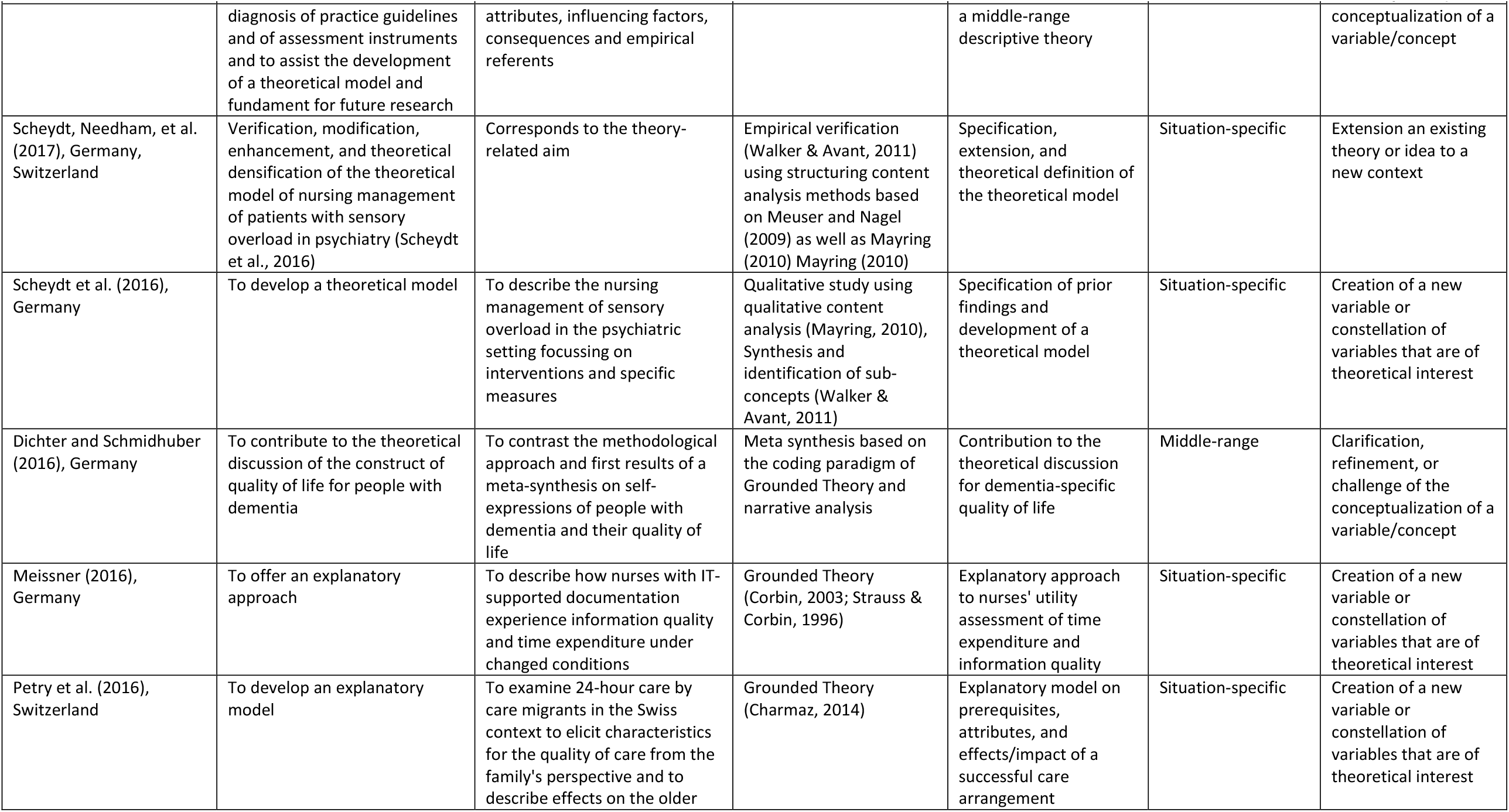
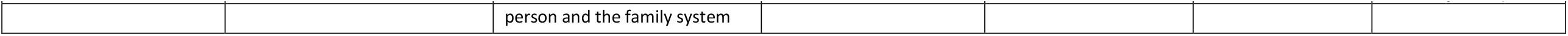
Data extraction of identified theoretical nursing research of the German-speaking area

### Theoretical contributions of nursing researchers from the German-speaking area of Europe

#### Creation of a new variable or constellation of variables (18)

Various authors created a new constellation of variables by means of a Grounded Theory. Wosko, Schnepp (28) spoke of a theoretical construct with three different trust patterns to explore how HIV/AIDS patients experience their everyday life. The constellation of variables by Oetting-Roß et al. (2018), also developed by means of a Grounded Theory consisted of four central concepts to form the process of being an ill child. Fleischmann et al. (2017) generated a theoretical construct of nurses’ experiences with general practitioners visits to the nursing home by conducting a Grounded Theory study. Meyer et al. (2017) developed a model of decision-making in complex home-like birth settings (Meyer et al., 2017) and Meissner (2016) conducted a Grounded Theory study to explain the benefit assessment by nurses of time expenditure and information quality (Meissner, 2016). The constructivist Grounded Theory study by Petry et al. (2016) resulted in an explanatory model on prerequisites, attributes and effects of a successful care arrangement (Petry et al., 2016) and the constructivist Grounded Theory study by Waldboth, Patch (26) lead to a constellation of four variables to go beyond description and generate an explanation of a process grounded in data.

Zumstein-Shaha et al. (2020) developed a theory including its concepts, their dimensions and subdimensions, to demonstrate the whole experience of the person living with cancer (Zumstein-Shaha et al., 2020). Roll and Bowers (2019) developed a theoretical framework to represent how families are involved in the development of support networks for their adult children with Down Syndrome and the strategies they use to achieve this goal. In the qualitative study by Bauer and Mayer (2019), the authors developed a conceptual order with focus on the strategies patients with multiple sclerosis use to deal with increasing immobilisation.

A meta-synthesis of qualitative studies about the experiences of relatives of people with dementia in acute hospitals (Burgstaller et al., 2018) resulted in a new interpretation on a theoretical level by means of a lines-of-argument synthesis. Using an embedded single case study design, Saladin et al. (2018) developed a central model of caring for a person during voluntary stopping of eating and drinking in a long-term care institution. To collate existing qualitative research into a comprehensive framework related to adult migrant patients’ experiences of health care, a meta-ethnography was conducted by (Luiking et al., 2019). The authors developed five factors that determine the migrant’s healthcare experience and evaluation, as well as antecedents and succedents (Luiking et al., 2019).

#### Clarification, refinement, or challenge of the conceptualization of a variable or a concept (18)

Huber et al. (2021) redefined the concept of patient-related complexity of nursing care for the setting of acute care hospitals. They advanced the concept by including the type, number and assessability of problems of patients and their relatives determining the extent of complexity (Huber et al., 2021).

Bauernschmidt and Dorschner (2018) clarified the designations used in the German-speaking nursing literature for persons in the close social environment of people receiving nursing care by discussing their suitability for nursing language use (Bauernschmidt & Dorschner, 2018).

Schirghuber and Schrems (2018) provided the basis for a conceptual definition to develop an evidence-based nursing diagnosis on the phenomena of boundedness and bedriddenness through operationalization (63).

Dandliker and Kolbe (2017) also aimed to develop a nursing diagnosis by exploring the concept of withdrawal phenomena and by analysing its defining characteristics, related factors, and risk factors. Scheydt, Staub, et al. (2017) analysed the meaning and inner structure of sensory overload in the psychiatric context for the development of a nursing diagnosis on the basis of a theoretical model (Scheydt, Staub, et al., 2017). Balaam et al. (2017) explored the concept of migrant women and proposed an operational definition for the use in research, policy, and targeted health service delivery. Grace and Zumstein-Shaha (2020) parsed out nursing and science as separate concepts and synthesized a definition of nursing science using the principle of Ockham’s razor, the law of parsimony in explanations (Grace & Zumstein-Shaha, 2020).

Dichter and Schmidhuber (2016) identified 19 dementia-specific quality of life instruments. Their aim was to contribute to the theoretical discussion of the construct of quality of life for people with dementia by means of a meta-synthesis (82).

#### Identification of mechanisms of intervening processes (18)

Kohler et al. (2020) developed a programme theory as basis for a theory-driven evaluation of a complex intervention for promoting continence for people who have suffered a stroke by making assumptions about the mechanisms of intervening processes (Kohler et al., 2020). In preparation for this, they previously conducted a qualitative study (Kohler et al., 2018) to consider the lived experience of affected persons. These findings served as indications for the development of the complex intervention.

Krieger et al. (2017) developed a complex intervention programme to support caregivers of patients who have suffered a stroke. It consists of five conceptual building blocks: content, human resources, personalised approach, timing and setting (Krieger et al., 2017).

In the context of dementia care, Adlbrecht et al. (2018) presented a programme theory of an existing care concept of an Austrian dementia special care unit and reflected on the methodology used for its development. The programme theory formed the basis for its theory-based evaluation. It consists of an action model as well as a change model, which explicates the mechanisms through which outcomes are expected to be achieved (Adlbrecht et al., 2018).

#### Extension of an existing theory or idea (18)

Mayer, Breuer (33) revised the conceptual model of medication self-management by Bailey et al. (2013) on the basis of a secondary qualitative data analysis. The revised model contains an expansion of the content, a differentiation of the individual steps, a reorganization of the steps and thus an expanded understanding of a more complex process logic. Gerlach and Schupp (2018) extended the theory of awareness of homosexuality (Plummer, 1973) by means of a Grounded Theory. The group of Scheydt, Needham, et al. (2017) further developed their own work of nursing care of patients with sensory overload in the psychiatric setting to empirically verify, modify, and enhance their existing framework (Scheydt, Needham, et al., 2017). Bohner (2017) examined the applicability of Meleis (2010) Transition Theory in Swiss nursing practice and research by evaluating the conceptual strengths and weaknesses by a theory description, critical reflection, and theory evaluation (66).

#### Synthesis of multiple theories into a unified framework (18)

Peters et al. (2020) conducted a theory synthesis by analysing existing concepts, theories and preferences of women to midwifery care. They developed a hierarchical model of the process of midwifery practice based on eleven identified grand or middle-range theories (Peters et al., 2020). Another theory synthesis was conducted to link illness-related emotional distress with symptom self-management in chronic respiratory disease. Schmid-Mohler et al. (2019) identified twelve relevant models and synthesized them into an overarching conceptual model, that describes the relationship between symptom perception, symptom self-management and outcomes (Schmid-Mohler et al., 2019).

## DISCUSSION

We provided an overview of the landscape of theoretical nursing research in the German-speaking area of Europe of the last years. We found that two percent of articles from nursing journals with at least one affiliation of an author from a German-speaking country explicitly pursued a theoretical aim. Most theoretical research followed an inductive and theory-building approach and mainly referred to Grounded Theory as methodology and concept analysis as methods. Theoretical results were mainly descriptive, situation-specific and presented as models. Illness experience and self-management were the most prevalent among the addressed phenomena.

Our review corroborates the thesis of Moers et al. (2011) regarding the sparse theory development in nursing in the German-speaking area, which can be maintained for the time period of 2016-2021 (as of March 29). This observation pertains not only to the quantitative amount of theoretical research, but also to the overall contribution to our discipline’s body of knowledge on a metatheoretical level, which we, however, can only assess approximately based on this work. Nonetheless, a first picture emerged which also reveals the diversity of theoretical nursing research in the German-speaking countries of Europe, which we would like to emphasize and give importance to at this point. However, the more appropriate question to ask is thus not whether we theorize or not, but *how* we theorize.

Moers et al. (2011) outlined implications and pending tasks for theory development. What they considered essential was the theoretical generalization of research findings. At that time, initial meta-analyses on living with chronic diseases had shown that the available findings, which originated predominantly from qualitative, and occasionally also from quantitative research, resembled a colorful patchwork without a coherent pattern.

Schaeffer (2009) demanded at that time to discuss the heterogenous research in a generalizing way. In the context of this mapping review, we also came to this finding, as the proportion of metatheoretical research, such as meta-synthesis or theory-synthesis, was small, and others consisted mostly of qualitative studies that resulted in situation-specific theories with little reference to the existing body of theory. Often, nonspecific designs and analysis methods were selected that were generally not oriented toward theory development. However, Grounded Theory methodology was prevalent, probably due to its well-structured and explicitly described approach. Also, the large proportion of concept analyses must be emphasized, while it remains unclear how many of these are related back to theory and how they relate to each other. This variety of incoherent research, however, fits well the aforementioned patchwork analogy (Schaeffer, 2009). Therefore, there is still a need for a revival of inductive theory discourses so that different health and nursing science theory discourses can be consolidated (Schaeffer, 2009). Another implication outlined by Moers et al. (2011) for future theory development was to strengthen the patient perspective and to consider new aspects, such as participation and self-management. Accordingly, there was a lack of research on the patient perspective on illness experience and coping actions, which was especially discussed against the background of creating a knowledge base for theoretically supporting intervention development. However, our review identified a positive development in this regard, with half of the studies focusing on the illness experience or self-management. We furthermore identified studies on programme theories, albeit on a small scale, which provide the theoretical foundation for the development and evaluation of interventions for patients and therefore represent another theoretical aspect of the patient perspective and the promotion of self-management.

Finally, Moers et al. (2011) have pointed out the need for the further and new development of existing theoretical work that respond to the changing demographic, epidemiological, social, and structural conditions of society and healthcare in the German-speaking area of Europe. Here again, based on the results of this focused mapping review, we identified a persistent need, as mainly inductive theory-building work was identified but less theory-testing or modifying efforts. However, existing theories are a valuable resource and should be used to build better ones. Theory-building should be dynamic, as the process of theorizing never ends. To that end, we agree with Turner (87), who cautioned that researchers shall not become “monks copying and reciting passages from the sacred texts” (Turner, 1990).

It is interesting, however, that the theoretical generalization of research findings as well as the further development of theories, so the actual processes of theory construction, still seem to be an issue today, although these problems have been discussed and repeatedly demanded by the scientific community for more than twenty years now in the German-speaking area of Europe. The question arises whether the sparse theory-development is primarily caused by researchers being “too busy to think” (Moers et al., 2011) due to the constraints and trends in the research system (Bartholomeyczik, 2018; Brandenburg, 2019; Dorschner, 2022; Moers et al., 2011) and because critical thinking is not promoted enough in the scientific community (Bartholomeyczik, 2018), or whether we might need to learn to think theoretically and to theorize (again) in the first place. Theorizing is profoundly communal in nature and following it will only succeed if it is deeply rooted in a community of scholars and a general culture of theorizing (Swedberg, 2011).

There have been discussions for many years, primarily in the field of sociology, but also in the context of the social sciences, about whether the majority of researchers may never have properly learned how to theorize (Swedberg, 2016). One potential influence may be the increasing focus on theories as an end product and less on the process of theorizing itself. Swedberg (2011) speaks in this sense of the scientific enterprise as consisting of three elements: 1) theorizing, 2) theory and 3) testing of theory. According to him only the second and the third elements were properly attended to over the last years, whereas the first element of theorizing was largely ignored by social scientists. The neglect of theorizing, and the related overemphasis on theory, is related to the tendency in today’s social science to leave aside the goal of discovery and instead focus on justification. This is also in line with the fact that the scientific developments in the last decades were centered around developing research methods and introducing empirical research. Theorizing, as the practical skill of constructing and handling theories, however, has not developed in a similar manner. This led to social sciences with highly developed methods but underdeveloped skills in theorizing (Swedberg, 2014), which can also be observed in nursing science. Regarding our sample the description of the process(es) of theorizing was in most cases limited to naming or referring to standardized methods and the reduction of the procedure to systematized steps. Whether this is to be considered a lack of focus on theorizing, or plainly the way of reporting, remains open at this point.

One relevant factor leading to this neglect is that the process of theorizing it is not taught effectively (Mayer, 2019). While traditional lecturing combined with classroom discussions may be the common way to teach theory, something different is needed when it comes to teaching theorizing. Learning to theorize is similar to learning methods, you cannot learn by reading about it but by actively practicing to think theoretically, and by using or constructing theories (Swedberg, 2012). A challenge lays in the path of theorizing which is a creative process that cannot necessarily be derived systematically like research methods (Mayer, 2019). Theorizing demands learning by doing (Mayer, 2019; Swedberg, 2019) and constant repetition of thought experiments until a creative theorizing culture and attitude emerges to think about the world and its phenomena in a theoretical manner (Jaccard & Jacoby, 2020). This theoretical culture should not only be promoted in teaching, but also in the scientific community itself. According to Krause (2016), theorizing happens in the interstices, such as in designing a project or research methods, in journal clubs for discussing papers, in libraries and coffee-shops. In order to create intellectual space for practices of theorizing, we should spell out the many different ways in which theorizing can contribute to knowledge production and explanation (Krause, 2016).

## Limitations of the study

This focused mapping review and synthesis aimed to provide an overview on theoretical nursing research landscape in the German-speaking countries of Europe. Nevertheless, this type of review has inherent limitations regarding its scope. Our descriptive “map” provides a snapshot, that is contextual and temporal and reflects only the published material in the included journals. However, as an impression of the state of theoretical nursing research in the German-speaking area of Europe, our review provided insights in theoretical activities over the last years. A further potential limitation concerns the focus on journal publications for the identification of theoretical research in our discipline. One could argue that theoretical work is rather published as a monograph due to their depth and extent, as already noticed by Bartholomeyczik (2018) that contributions as books often go unnoticed. Therefore, publications in journals should only be viewed with caution as an indicator of the status of theory development. Another limitation concerns our understanding of theoretical nursing research. The starting point for our analysis was to make decisions about whether an article was “theoretical” or not. This proved more of a challenge than we had anticipated, as judging articles for being theoretical by predefined categories was difficult. Nevertheless, we were able to work discursively on an inclusion criterion that still in the end was not always clear. Therefore, there is a high probability that we did not recognize or deliberately excluded work, although it might have been relevant for the scope of this mapping review. While with this work we can provide an overview of the theoretical characteristics of the included research articles, this focused mapping approach does not permit to give an overview of the process of theorizing.

## CONCLUSION

Since an imbalance between empirical and theoretical nursing inquiry seems to be an issue in other nursing science communities as well(Grace et al., 2016; Roy, 2018), we assume that the significance of the results of this study goes beyond the cultural context of the German-speaking region of Europe.

More attention should be drawn to find a routine in thinking theoretically in nursing research. To this end, more efforts should be made on how we can work on theorizing rather than focusing exclusively on the theories that are available to us. Nursing researchers should be properly educated in theoretical thinking as well as in methodology of theory construction. Finally, we should actively revive theorizing, e.g. through scientific symposia on theory building, research projects on working theoretically and by living a culture in which theorizing is demanded and encouraged. Nursing professors have a crucial role to defy the demands of the current research mainstream and provide appropriate structures to learn to theorize, because “to think at all is to theorize” (Coleridge, 1812).

## Data Availability Statement

All data generated for the purpose of this study are part of the manuscript and supplementary material.

## Supporting information

List of nursing journals

